# Assessing the quality, readability and reliability of online information on COVID-19: an infoveillance observational study

**DOI:** 10.1101/2020.05.30.20117614

**Authors:** Paulo Cardoso Lins-Filho, Thuanny Silva de Macêdo, Andressa Kelly Alves Ferreira, Maria Cecília Freire de Melo, Millena Mirella Silva de Araújo, Jaciel Leandro de Melo Freitas, Thaise Urbano Caldas, Arnaldo de França Caldas

**Author notes:** **Corresponding Author:** Arnaldo de França Caldas Júnior Estrada de Aldeia, Km 13, Prive Portal de Aldeia, Aldeia, Camaragibe, Pernambuco, Brasil.

## Abstract

**Objective:** This study aimed to assess the quality, reliability and readability of internet-based information on COVID-19 available on Brazil’ most used search engines.

**Methods:** A total of 68 websites were selected through Google, Bing, and Yahoo. The websites content quality and reliability were evaluated using the DISCERN questionnaire, the Journal of American Medical Association (JAMA) benchmark criteria, and the presence of the Health on Net (HON) certification. Readability was assessed by the Flesch Reading Ease adapted to Brazilian Portuguese (FRE-BP).

**Results:** The web contents were considered moderate to low quality according to DISCERN and JAMA mean scores. Most of the sample presented very difficult reading levels and only 7.4% displayed HON certification. Websites of Governmental and health-related authorship nature showed lower JAMA mean scores and quality and readability measures did not correlate to the webpages content type.

**Conclusion:** COVID-19 related contents available online were considered of low to moderate quality and not accessible.

## Introduction

Health care is rapidly transitioning from a paternalistic approach to a person-centered model. This process aims to improve health outcomes by building a shared decision-making process between healthcare professionals and patients, characterized by the greater involvement of people in resolutions and actions concerning their own health (Lee et al. 2018; Petersen et al. 2019). However, the effectiveness of this new model can be hindered by a considerable number of barriers, such as low education, inadequate access to knowledge and social and economic deprivation (Lee et al. 2017).

The internet offers a large amount of information, although the quality of health and sanitary information offered is highly variable, ranging from scientific and evidence-based data to home remedies or information of very questionable origin that can be dangerous to health (Eysenbach et al. 2002). The biggest barrier on the internet is not the difficulty of finding health care information, but identifying those that are valid and reliable (Berland et al. 2001; Lopez-Jornet and Camacho-Alonso 2009; Lopez-Jornet and Camacho-Alonso 2010; Passos et al. 2020).

During public health emergencies, people must be aware about the health risks they face, and what measures can be taken to protect their health and lives. Reliable information provided early, often, and in accessible language standards, enables individuals to make choices and act to protect themselves, their families and communities from health hazards (WHO 2017). Fake news and misinformation concerning health on the internet represents a threat to global health (Carrieri et al. 2019). The World Health Organization (WHO) warned that the COVID-19 outbreak had been accompanied by a massive abundance of information, some of which was accurate and some of which was not, which made it difficult for people to find reliable sources and trustworthy information when they needed it (Kouzy et al. 2020; Pulido et al. 2020). The consequences of disinformation overload are the spread of uncertainty, fear, anxiety and racism. Thus, monitoring the quality of information available to the population is of great importance to control the spread of the disease itself and to mitigate its socioeconomic impacts (Hua and Shaw 2020). Therefore, the WHO is dedicating tremendous efforts aimed at providing evidence-based information and advice to the population through its social media channels and a new information platform called WHO Information Network for Epidemics (Zarocostas 2020).

Online-available information has been increasingly employed as a surrogate tool for estimating epidemiology (Cervellin et al. 2017). Web-based sources are been used in the analysis, detection, and forecasting of diseases and epidemics, and in predicting human behavior toward several health topics. In this context, infoveillance studies have become an integral part of health sciences that focuses on scanning the Internet for user-contributed health-related content, aiming to improve public health, measuring and predicting the quality of health information on the Web (Eysenbach et al. 2009).

Several studies have already assessed the quality of the information available on the internet related to different health conditions (Cuan-Baltazar et al. 2020; Jo et al. 2018; Lee et al. 2017; Passos et al. 2020; Priyanka et al. 2018), however, there is no evidence about the quality of COVID-19 contents available in Brazil. This study aimed to assess the reliability, readability and quality of COVID-19 related information retrieved from Brazilian websites.

## Materials and Methods

The search strategy was designed with regard to the relevance of terms employed by the internet users. A query was performed on Google Trends to confirm the link of Brazilian Portuguese words to COVID-19 issues. Google Trends enables researchers to study the trends and patterns of Google search queries (Effenberger et al. 2020). The search term “coronavírus” held the most popularity among internet users in Brazil, in April 2020.

Sites on the internet were identified using the three most accessed search engines by internet users in Brazil: Google www.google.com), Bing (www.bing.com) and Yahoo www.yahoo.com), respectively, 97.59%, 1.2% and 1.04% of accesses in April 2020 (Statcounter 2020). In April 2020, the searches were history of each browser. The first 100 consecutive sites in each search were visited and classified. The search was not restricted in terms of file format or domain. The search was limited to the Portuguese language. Duplicate sites were excluded, as were non-operative sites or sites with denied direct access through password requirements, book review sites, or sites offering journal abstracts, and those sites that did not offer information on COVID-19. Websites that could be modified by the general population were also not considered in this investigation.

The quality of website information was assessed by four evaluators, who were previously trained in the analysis tools used. Concerning the scientific accuracy and reliability of websites information, WHO official reports and technical guidelines were used as standards. The websites that were divergently qualified by the examiners were reassessed to the achievement of a consensus score. As this was a study of published information and involved no participants, no ethics approval was required. In order to avoid any changes that may be made to the eligible websites during the period of analysis, the sites were assessed in the same day by the evaluators.

The sites were classified in terms of affiliation as commercial, news portal, non-profit organization, university or health center and government. The type of content was classified as corresponding to medical facts, human experiences of interest, questions and answers and socioeconomic related content.

The quality of information of the selected websites was assessed using criteria of the Journal of the American Medical Association (JAMA) benchmarks (Silberg et al. 1997). These are a display of authorship of medical content, display of attribution or references, display of currency (date of update), and disclosure of ownership, sponsorship, advertising policies or conflicts of interest. This tool lets the reader easily decide if the site has the basic components like transparency and reliability. For each fulfilled criterion, 1 point was given, with a total score ranging from 0 to 4.

The DISCERN instrument (Discern) is a valid and reliable tool to evaluate health information. It is the first standardized quality index and was created by the Division of Public Health and Primary Health Care at Oxford University, London. The instrument comprises 16 questions, each representing a different quality criterion. The DISCERN questions are organized into three sections as follows: Questions 1–8 performed using computers connected to the internet, previously set up by clearing the cookies and search address the reliability of the publication and help users to decide whether it can be trusted as a source of information relating to treatment choice. Questions 9–15 address specific details of the information relating to treatment alternatives. In this context, questions 9–11 refer to the active treatments described in the publication (possibly including self-care), while the options without treatment are addressed separately in question 12. In turn, question 16 corresponds to the global quality assessment at the end of the instrument. Each question is scored on a scale of 1–5 (where 1, the publication is poor; and 5, the publication is of good quality). In the present study, only the first section of the questionnaire was used for reliability assessment.

The readability (RE) of the websites was assessed by the Flesch Reading Ease adapted to Brazilian Portuguese (FRE) (Lee et al. 2017). This method classifies the readability of a text on a scale from 0 (very difficult) to 100 (very easy) based on a calculation that considers the number of syllables per word and words per sentence. The adapted formula is given by the following equation: RE = 248.835 − (1.015 × ASL) − (84.6 × ASW). Where: ASL = mean number of words per sentence; ASW = mean number of syllables per word.

Those metrics were calculated using the online tool Readable.io (Readable.io, Bolney, England) (Readable.io) through the information of the respective Uniform Resource Locator (URL) of each website. All analyses were performed based on the overall written content downloaded from these links. The reading difficulty of a text is presented according to the following scores: very easy (75–100), easy (50–75), difficult (25–50), and very difficult (0–25).

The existence of the Health on the Net (HON) Foundation seal was also recorded. HON is a code of conduct for medical and healthcare sites, defining a series of norms allowing users to know the source and the purpose of the medical information presented. The HON contemplates compliance with the following eight basic criteria: 1. authorship; 2. complementarity; 3. privacy; 4. attribution, references and currency; 5. justifiability; 6. author transparency; 7. sponsor transparency (financial disclosure); and 8. honesty in advertising policy. The website may display the HON code seal if they agree to comply with the standards listed, and they are subjected to random audits for compliance(HON).

Data were submitted to statistical analysis, all tests were applied considering an error of 5% and the confidence interval of 95%, and the analyzes were carried out using SPSS software version 23.0 (SPSS Inc. Chicago, IL, USA). Descriptive analysis was performed to characterize the Web pages selected for the study. Although the hypothesis of normal distribution of data was not confirmed by the Kolmogorov-Smirnov test, the statistical analysis was performed by the application of nonparametric tests. The correlations between distinct measures were demonstrated by the Spearman rank correlation coefficients. Distinct websites according to the type of content were compared by Kruskal-Wallis test. Mann-Whitney U test was employed to assess the differences between the natures of websites, for this test, the affiliation criteria used in data collection were dichotomized into governmental and health-related authorship nature (grouping university or health center and government affiliation) and nongovernmental nor health-related authorship nature (grouping news portals, commercial sites and non-profit organization affiliation)(Lee et al. 2017).

## Results

Over 2.5 billion results were retrieved from the search engines. Of the 300 webpages assessed, 68 fulfilled the inclusion and exclusion criteria. The search retrieval flow diagram is presented in Figure 1.

**Fig. 1.**
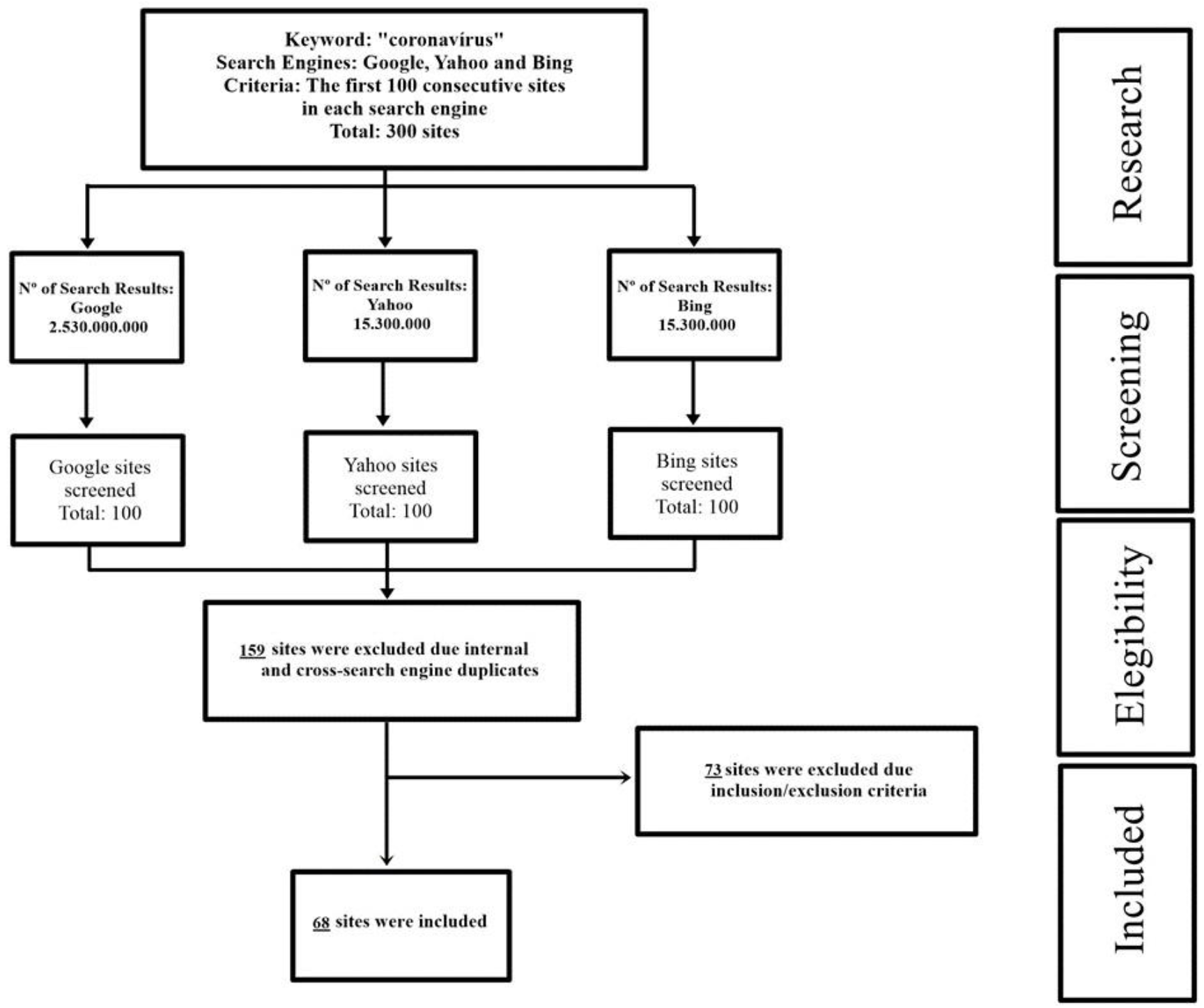
Search retrieval flow diagram

According to affiliation most websites were from news portals (51.5%), followed by government (29.4%), commercial sites (13.2%), university or health center (4.4%) and non-profit organization (1.5%). Considering the type of content, the majority of the sites displayed medical facts (88.2%) followed by socioeconomic related content (5.9%), questions and answers (4.4%) and human experiences of interest (1.5%).

None of the evaluated websites met all four criteria of JAMA benchmarks, 35.3% had a single criterion, 30.9% did not include any criteria, 23.5% had 2 criteria and 10.3% presented 3 criteria. Only 7.4% of the sites had HON certification. The DISCERN instrument identified that 75% of the websites had moderate reliability, 17.6% showed high reliability and 7.4% low reliability. For the Flesch Index, over half of the sample were classified as very difficult (57.4%), while 41.2% were considered difficult. Only 1 website (1.5%) was classified as easy and none as very easy. Sample means of JAMA, DISCERN and FREBP scores are shown in table 1.

**Table 1.** Descriptive statistics of the scores of DISCERN, the Journal of American Medical Association benchmark, and Flesch Reading Ease

|  | DISCERN | JAMA<sup>a</sup> | FRE-BP<sup>b</sup> |
| --- | --- | --- | --- |
| Mean (SD) | 1.90 (0.493) | 1.13 (0.976) | 22.32 (12.325) |
| Median | 2.00 | 1.00 | 23.00 |
| Minimum | 1.00 | 0.00 | 1.00 |
| Maximum | 3.00 | 3.00 | 72.00 |

<sup>a</sup>JAMA: Journal of American Medical Association.
<sup>b</sup>FRE-BP: Flesch Reading Ease adapted to Brazilian Portuguese.

The correlation between distinct measures assessed through the instruments was analyzed. Spearman rank correlation coefficients showed a positive significant correlation between JAMA and Discern scores (p<0.001), and between HON certification presence and JAMA (p = 0.006) and Discern < scores, as shown in table 2

**Table 2.** Correlations between distinct quality measures

| Quality measures |  | Correlation coefficient (p-value) |  |
| --- | --- | --- | --- |
| JAMA<sup>a</sup> | 0.464* (<0.001)<sup>ab</sup> | -0.045 (0.716)<sup>ac</sup> | 0.329* (0.006)<sup>ad</sup> |
| DISCERN<sup>b</sup> | 0.464* (<0.001)<sup>ab</sup> | -0.182 (0.138)<sup>bc</sup> | 0.531* (<0.001)<sup>bd</sup> |
| FRE-BP<sup>c</sup> | -0.045 (0.716)<sup>ac</sup> | -0.182 (0.138)<sup>bc</sup> | -0.018 (0.882)<sup>cd</sup> |
| HON<sup>d</sup> | 0.329* (0.006)<sup>ad</sup> | 0.531* (<0.001)<sup>bd</sup> | -0.018 (0.882)<sup>cd</sup> |

<sup>a</sup>JAMA: Journal of American Medical Association.
<sup>c</sup>FRE-BP: Flesch Reading Ease adapted to Brazilian Portuguese.
<sup>d</sup>HON: Health on Net.
Superscript letters indicate correlation groups
\*Significant statistical difference between the groups (Spearman correlation coefficients, $p < 0.05$ ).

No significant differences were observed among the mean scores of DICERN, JAMA and FRE-BP according with the type of websites content (table 3). As the human experience of interest type of content represents a single occurrence in the sample it was disregarded in this step of the statistical analyzes.

**Table 3.** Descriptive statistics of different websites type of content for DISCERN, the Journal of American Medical Association benchmark, and Flesch Reading Ease adapted to Brazilian Portuguese

| Site content | DISCERN | JAMA<sup>a</sup> | FRE-BP<sup>b</sup> |
| --- | --- | --- | --- |
| Medical facts (n=60) |  |  |  |
| Mean <sup>(SD)</sup> | 1.90 <sup>(0.511)</sup> | 1.10 <sup>(0.986)</sup> | 22.38 <sup>(12.806)</sup> |
| Median | 2.00 | 1.00 | 23.00 |
| Minimum | 1.00 | 0.00 | 1 |
| Maximum | 3.00 | 3.00 | 72 |
| Questions and answers (n=3) |  |  |  |
| Mean <sup>(SD)</sup> | 1.67 <sup>(0.577)</sup> | 1.33 <sup>(1.115)</sup> | 25.00 <sup>(2.646)</sup> |
| Median | 2.00 | 2.00 | 24.00 |
| Minimum | 1.00 | 0.00 | 23.00 |
| Maximum | 2.00 | 2.00 | 28.00 |
| Socioeconomics (n=4) |  |  |  |
| Mean <sup>(SD)</sup> | 2.00 <sup>(0.000)</sup> | 1.50 <sup>(1.000)</sup> | 23.00 <sup>(9.274)</sup> |
| Median | 2.00 | 2.00 | 23.50 |
| Minimum | 2.00 | 0.00 | 14.00 |
| Maximum | 2.00 | 2.00 | 31.00 |
| p-value* | 0.818 | 0.780 | 0.555 |

aJAMA: Journal of American Medical Association.
bFRE-BP: Flesch Reading Ease adapted to Brazilian Portuguese.
\*Kruskal-Wallis test (p<0.05).

The mean JAMA scores were different according to the dichotomized affiliation categorization. Higher mean was observed in nongovernmental nor health-related authorship nature. For DISCERN and FRE-BP mean scores no significant statistical difference was observed, as shown in table 4.

**Table 4.** Descriptive statistics for both affiliation website groups for DISCERN, the Journal of American Medical Association benchmark, and Flesch Reading Ease adapted to Brazilian Portuguese

| Quality scores | Websites |  |  |  |  |  |  |  | p-value |
| --- | --- | --- | --- | --- | --- | --- | --- | --- | --- |
|  | Governmental and health-related nature<br>(n=23) |  |  |  | Nongovernmental nor health-related nature<br>(n=45) |  |  |  |  |
|  | Mean<br>(SD) | Median | Minimum | Maximum | Mean<br>(SD) | Median | Minimum | Maximum |  |
| DISCERN | 1.87<br>(0.548) | 2 | 1 | 3 | 1.91<br>(0.468) | 2 | 1 | 3 | 0.691 |
| JAMA<sup>a</sup> | 0.70<br>(1.063) | 0 | 0 | 3 | 1.36<br>(0.857) | 1 | 0 | 3 | 0.002* |
| FRE-BP<sup>b</sup> | 18.87<br>(9.725) | 22 | 1 | 40 | 24.09<br>(13.213) | 25 | 3 | 72 | 0.083 |

<sup>a</sup>JAMA: Journal of American Medical Association.
<sup>b</sup>FRE-BP: Flesch Reading Ease adapted to Brazilian Portuguese.
\*Significant statistical difference between the groups (Mann-Whitney U test, $P < 0.05$ ).

## Discussion

The internet has great potential for spreading health information (Berland et al. 2001). Whatever the communication channel used, it is known that its content is able to influence the decisions of an individual about his health, including changes in lifestyle (Afshin et al. 2016). Web-based information influences how patients comply with advices, clinical diagnoses, and treatment regimens recommended by health professionals (Lu et al. 2018; Lu and Zhang 2019).Treatment adherence and compliance relies on trust and good professional-patient communication (Wahl et al. 2005). In this context, internet-based new technologies are gaining growing global attention and becoming increasingly available for predicting, preventing and monitoring emerging infectious diseases, such as COVID-19 (Effenberger et al. 2020; Yang et al. 2020). However, misinformation spread through the internet can hinder the communication of health entities and professionals with the general population (Lu and Zhang 2019) and so reduce adherence to the confrontations proposed to contain the pandemic, as social distancing (Farooq et al. 2020). Research on the role of internet content, social media messages and dominant discourses that are communicated to the public is an emerging topic of public health interest in scientific work that requires further investigation (Pulido et al. 2020).

Despite the harm that misinformation may pose, especially during a pandemic in which the population’s reaction to health measures imposed by governments is of crucial importance to combat the spread of the disease (US Medicine Institute 2002), scientific evidence on the topic is scarce. Besides WHO efforts to monitor and improve the quality of information available online on this subject (Hua and Shaw 2020), the few available evidence warns of the low quality and reliability of data (Abd-Alrazaq et al. 2020; Febres-Cordero et al. 2018; Kouzy et al. 2020). These works are, however, limited to social media content analysis and lacking standard parameters to data evaluation. In the present study, for all parameters used in the data analysis, the quality of information ranged from low to moderate.

The HON seal was displayed in only 7.5% of the sample. Other studies carried on Brazilian websites revealed even a small number of sites with this certification (Lee et al. 2017; Passos et al. 2020). The absence of this seal on sites of important institutions indicates that concern for certifying the quality of information in the internet is still scarce. Another possible cause for low seal adhesion is the fact that the annual review required for seal maintenance is not free (Passos et al. 2020).

Besides the quality of information, the amount of it provided for an individual is also a concern. Previous research suggests that the vast amount of available information can be confusing, potentially resulting in over-concern and information overload (Farooq et al. 2020). Information overload is being associated to mental health problems during COVID-19 outbreak. These findings inspire the need for greater government involvement to prevent information overload while facing a public health emergency (Gao et al. 2020). However, government represents only 29.4% of the sources of information, while news portals represent 54.5% of the website’s affiliation. In addition, governmental and health-related sources of information showed lower JAMA mean scores when compared to nongovernmental nor health-related sources. This demonstrates the need for better articulation between health entities and government to stand out as the main provider of reliable content, directing users to good quality sources and avoiding information overload.

According to the readability scores, the websites were considered difficult and very difficult for most of population. In addition to this finding it is relevant to consider the low level of health literacy, which is the degree to which people have the capacity to understand health information, reported for Brazilian Portuguese speakers (Batista et al. 2018). This may result in a communication gap for laypersons.

In addition to ensure the quality and reliability of information, it is important that these quality contents are presented in a comprehensible and accessible manner (Miguens-Vila et al. 2018). Studies reported a negative correlation of readability scores with JAMA (Sobota and Ozakinci 2015) and DISCERN (Lee et al. 2017) scores. These findings can be considered as exacerbating factors of the impact of the low quality of information on internet users, as it demonstrates that more accessible content is of even worse quality. In the present study such correlation was not found possibly due the growing concern about the quality of health-related information available online (Farooq et al. 2020), however, no positive correlation was found, which demonstrates the need for further efforts on improving the accessibility of high-quality health related information available online.

The internet and search engines are dynamic processes that constantly change. The sites evaluated in this investigation may not necessarily reflect the information available to patients at another time. This was a limitation for this investigation. However, the search engines used for the consultation represent 99.8% of the access of Brazilian internet users (Statcounter 2020). In addition, to cover a reasonable amount of data, the first 100 consecutive websites of each search engine were accessed.

Regarding the present sample of Brazilian websites, COVID-19 contents were considered of low to moderate quality and low readability based on the parameters adopted. This pattern just reasonably correlated with the nature of websites’ authorship. These findings indicate the need for further efforts on improving the quality of health-related content on internet. Health authorities might apply this evidence to measure the effect of the transmission of information on the population and define better risk communication strategies.

## Data Availability

The database is available.

## Compliance with Ethical Standards

Conflict of interest: The authors declare that they have no conflict of interest.

## Acknowledgments

The authors would like to thank the Coordenação de Aperfeiçoamento de Pessoal de Nível Superior(CAPES) for scholarship granting.

